# Federated Learning-Driven Collaborative Diagnostic System for Metastatic Breast Cancer

**DOI:** 10.1101/2023.10.20.23297323

**Authors:** William Gao, Dayong Wang, Yi Huang

## Abstract

Metastatic breast cancer is one of the leading causes of cancer mortality. While there has been progress in developing deep learning-driven diagnostic system for metastatic breast cancer based on histopathological images, it faces a major challenge in real-world application, i.e., how to improve generalizability of the diagnostic models for diverse patient populations and variations in sample and image processing across different geographic regions. Multi-institutional collaborative learning, which trains a single model using data from multiple institutions, can address the challenges of data inadequacy, lack of data diversity, and data quality. However, the current practice of direct medical data transfer among institutions faces increasing restrictions including patient privacy, intellectual property, data ownership and legal obligations. To enable multi-institutional collaborative learning for cancer diagnosis, we developed a federated learning-driven collaborative diagnostic system for metastatic breast cancer. This system preserves patient privacy by decoupling model training from the need for direct medical data transfer between institutions. Further, this study has demonstrated that the federate learning-driven system can improve diagnostic model accuracy and generalizability by leveraging information derived from diverse data sources across multiple institutions. This study has also shown that this collaborative diagnostic system has a strong potential of improving local diagnosis performance on lower quality images at a resource constrained institution. Our research indicated that federated learning-driven diagnostic system is a feasible and robust framework for multi-institutional collaborative development of diagnostic models for metastatic breast cancer. In addition to the benefits of improving diagnostic model generalizability for diverse patient populations, the collaborative diagnostic system presents a new opportunity to enable under-resourced healthcare institutions to leverage external data resources to improve local diagnosis performance while preserving patient privacy.

## 1. Introduction

Advances in Artificial Intelligence, especially in the field of deep learning, presents new opportunities for innovations in cancer diagnosis and treatment ^[1]^. However, real-world application of deep learning-based diagnostic system requires that such a system can perform diagnosis with high accuracy for a broad population with diverse demographic characteristics, accounting for regional differences in diagnostic practice and quality. A diagnostic model trained from the data of a specific institution tends to overfit its data at home institution and has a limited ability to be generalized to the new data from other institutions ^[2]^. Due to lack of generalizability to be applied to diverse patient populations, model training based on diverse data from multiple institutions becomes crucial for building accurate, precise, and generalizable diagnostic models for broader patient populations.

Although significant progress has been made to develop machine learning-based image analysis methods for metastatic breast cancer diagnosis ^[3, 4, 5, 6]^, real-world clinical application of these methods remains a major challenge. Particularly, lack of access to adequate, diverse, and high-quality data for model building is exacerbated in under-resourced healthcare institutions in many developing countries. First, these institutions face severe shortages of trained pathologists. For instance, the average number of pathologists per head of population is 1 to 1,000,000 in sub-Saharan regions, compared with the ratio of 1 pathologist to 15,000–20,000 in the US and UK ^[7]^. Second, many of the under-resourced healthcare institutions serve sparsely populated and remote regions. Third, diagnostic quality may be impacted by lack of resources and training ^[8]^. Therefore, for under-resourced healthcare institutions, internal data may be inadequate in size and quality for diagnostic model building. These institutions are in critical need to access and use high-quality external data for improving diagnostic model performance.

Multi-institutional collaborative learning, which trains a single model using data from multiple institutions, is a robust way to address the challenges of data inadequacy, lack of data diversity, and data quality. The current paradigm for multi-institutional collaboration is that the participating institutions physically transferred their data to a centralized location for model training and testing ^[9]^. While the centralized data sharing approach can increase the size of dataset for model training, the direct data transfer among healthcare institutions faces restrictions from multiple fronts, including patient privacy, intellectual property, data ownership. Among many countries, the United States Health Insurance Portability and Accountability Act (HIPAA) and the European General Data Protection Regulation (GDPR) regulate storage and exchange of personally identifiable data and health data ^[10, 11]^. Medical image data often cannot be shared outside the institutions of their origin due to patient privacy and confidentiality. Therefore, the centralized data sharing approach faces insurmountable difficulties in real-world applications, and therefore it is not feasible to scale this approach to international or global level.

Federated learning presents an innovative collaborative learning paradigm which enables models to be trained across multiple medical institutions without direct data transfer among them.

Federated learning decouples model training from the need for direct access to the raw training data, which significantly reduces the privacy and security risks ^[12. 13]^. Federated learning makes it possible to build global collaborative AI-driven systems for medical diagnosis, treatment, and disease monitoring ^[14]^. Further, such systems provide a crucial opportunity to help improve efficiency and quality of healthcare in remote, resource-constrained, or sparsely populated areas. Breast cancer patients in developing countries, especially sub-Saharan Africa, suffer from the highest mortality rates in the world due to lack of trained pathologists and consequent long diagnosis delays ^[15]^. While progress has been made to design AI-driven diagnostic systems for a single institution, the systems are limited to the data generated by the institution. However, a major challenge for under-resourced institutions is severe shortage of high-quality medical image data for building high-performing diagnostic models. This is further complicated by different patient populations and varying degrees of image quality. The diagnostic models trained on datasets procured from a single institution or institutions in a specific region may not perform as expected when applied to data from other institutions serving different patient populations. Federated learning provides an innovative framework for leveraging external demographically and geographically diverse data sources as well as incorporating local data to improve model accuracy and generalizability. The objective of this research is to develop a federated learning-driven collaborative diagnostic system for metastatic breast cancer by integrating knowledge from multiple institutions while preserving patient privacy.

## 2. Methods

### 2.1. Overall framework of federated learning-driven diagnostic system

Direct data sharing is the current approach for aggregating medical image data for diagnostic model training, which requires healthcare institutions to directly transfer their medical image data to a central server (Figure 1A). However, this approach is restricted by the laws and regulations on patient privacy, confidentiality, and data security ^[10, 11]^. To address this challenge, we propose a federated learning-driven collaborative diagnostic system for metastatic breast cancer. As shown in Figure 1B, the collaborating healthcare institutions do not directly transfer their medical image data to a central server. Instead, each institution trains a local copy of a diagnostic model and sends updates of model parameters to the central server without disclosing local medical data. The central server then aggregates the updates for the federated model and sends the newly updated model parameters to each of the collaborating institutions for further training or application. The federated learning-driven system substantially reduces the risk for patient privacy and confidentiality as compared with direct data transfer. Additionally, the communication cost of the federated learning-driven system is substantially lower than direct data transfer. As shown in Figure 1, the communication cost of the federated system of four institutions was 1280Mb for 10 iterations; in comparison, the cost for direct transfer of a set of 220 images was 306Gb.

**Figure 1.**
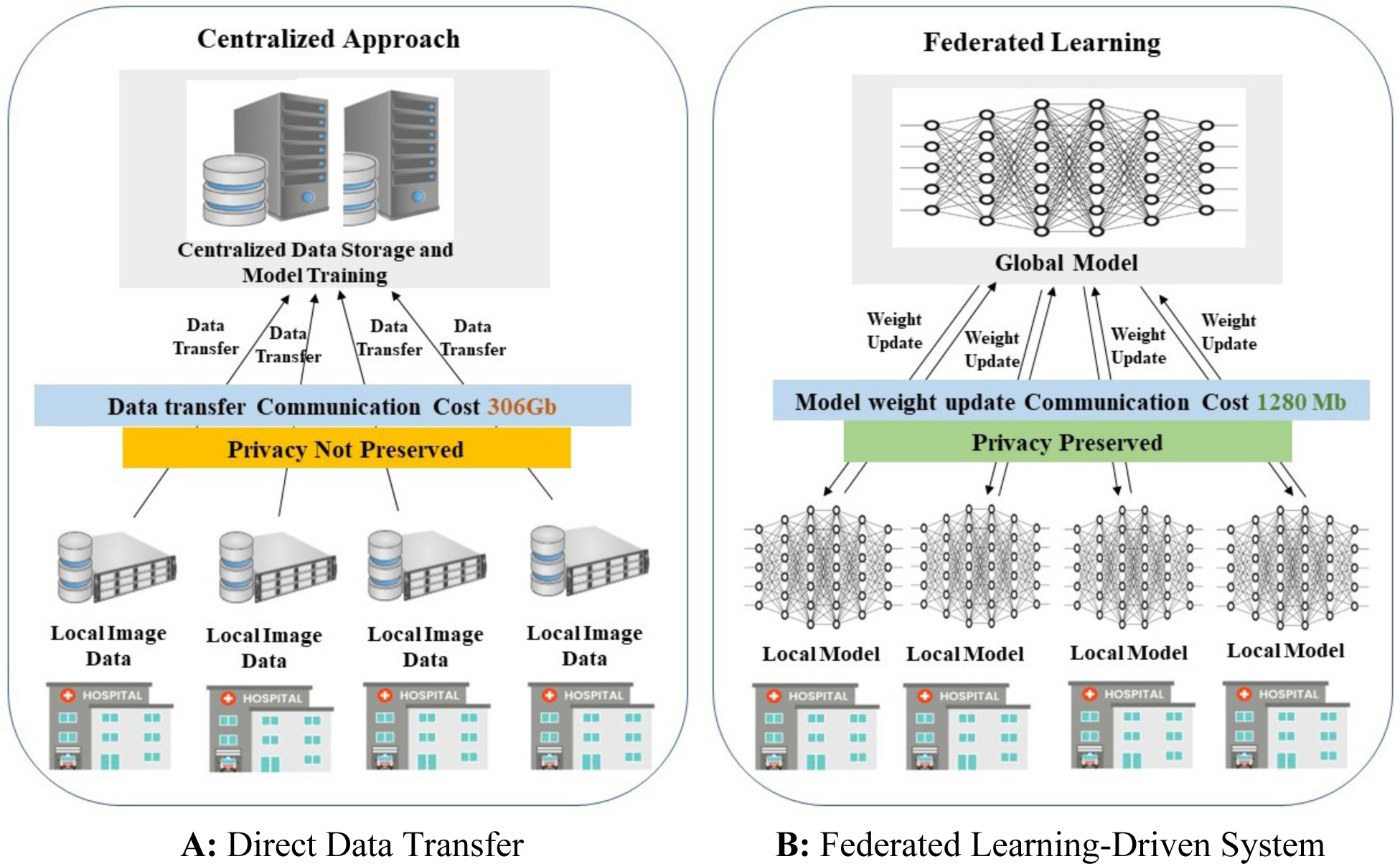
Overall framework of the federated learning-driven collaborative diagnostic system (B) for metastatic breast cancer, in comparison with the direct data sharing approach (A)

### 2.2. Histopathological image processing

Deep learning -based diagnosis for metastatic breast cancer is based on histopathological whole slide images (WSIs) of sentinel lymph nodes with a pathologist’s delineation of regions of metastatic cancer. The image normalization was conducted with the WSI Color Standardization procedure ^[16]^ to minimize potential variations in the color and intensity of H&E staining. Tissue areas within the normalized WSIs were identified and extracted using a threshold-based segmentation method ^[17]^. The mask images were generated for model training from the HSV representation transformed from the original RGB images. The WSIs were segmented into small patches which enable the deep learning model to be trained to recognize cancerous cells on a small scale, improving data volume and model training efficiency ^[18, 19]^. In the segmentation process, a large number of patches were extracted from each WSI and categorized as a positive tumor, negative tumor, and negative normal. A positive tumor patch was extracted from a tumor slide, containing cancerous regions; a negative tumor patch was from a tumor slide but did not contain a cancerous region; a negative normal patch was extracted from a noncancerous/normal slide. During patch extraction, masks were created using a lower and upper bound of pixel color. After reducing mask noise with the “opening” and “closing” morphology processes, the pixel values of each mask were used to categorize the patch.

### 2.3. Convolutional neural network (CNN) for single-institution diagnostic system

The local CNN-based diagnostic systems at individual institutions constitute the basic units of the federated learning-driven collaborative diagnostic system. MobileNetV2 was used for the single-institution diagnostic system. Mobilenetv2 is a CNN architecture that aims to perform computer vision tasks efficiently on mobile devices, by incorporating the Inverted Residual Structure and the Depthwise Separable Convolution to significantly reduce the model size and complexity ^[20]^.

The local CNN-based diagnostic system consists of two components: (1) training to build CNN-based diagnostic models; (2) testing for prediction on unseen WSIs (Figure 2). The training samples were WSIs with the corresponding ground truth image annotation indicating the delineation of regions of metastatic cancer. The diagnostic models were trained to discriminate between cancerous and noncancerous patches using a large number of small positive and negative patches randomly extracted from the set of training WSIs. We used five-fold cross-validation to evaluate classification performance. For an individual fold, each model ran through ten iterations or “epochs”. Each epoch allowed the model to reevaluate its weights to determine a more effective set of values. With a stochastic gradient descent (SGD) optimizer, the weights were retrained or optimized for the data with a 0.01 learning curve. Model accuracy was assessed for the training and validation set at each epoch. The model performance was assessed with independent (unseen) WSI image data. The unseen WSIs were pre-processed and segmented as described above. The diagnostic models were used to discriminate the cancerous vs. non-cancerous patches. With the patch-level classification, a tumor probability heatmap was generated for each WSI to indicate the probability of each pixel being cancerous.

**Figure 2.**
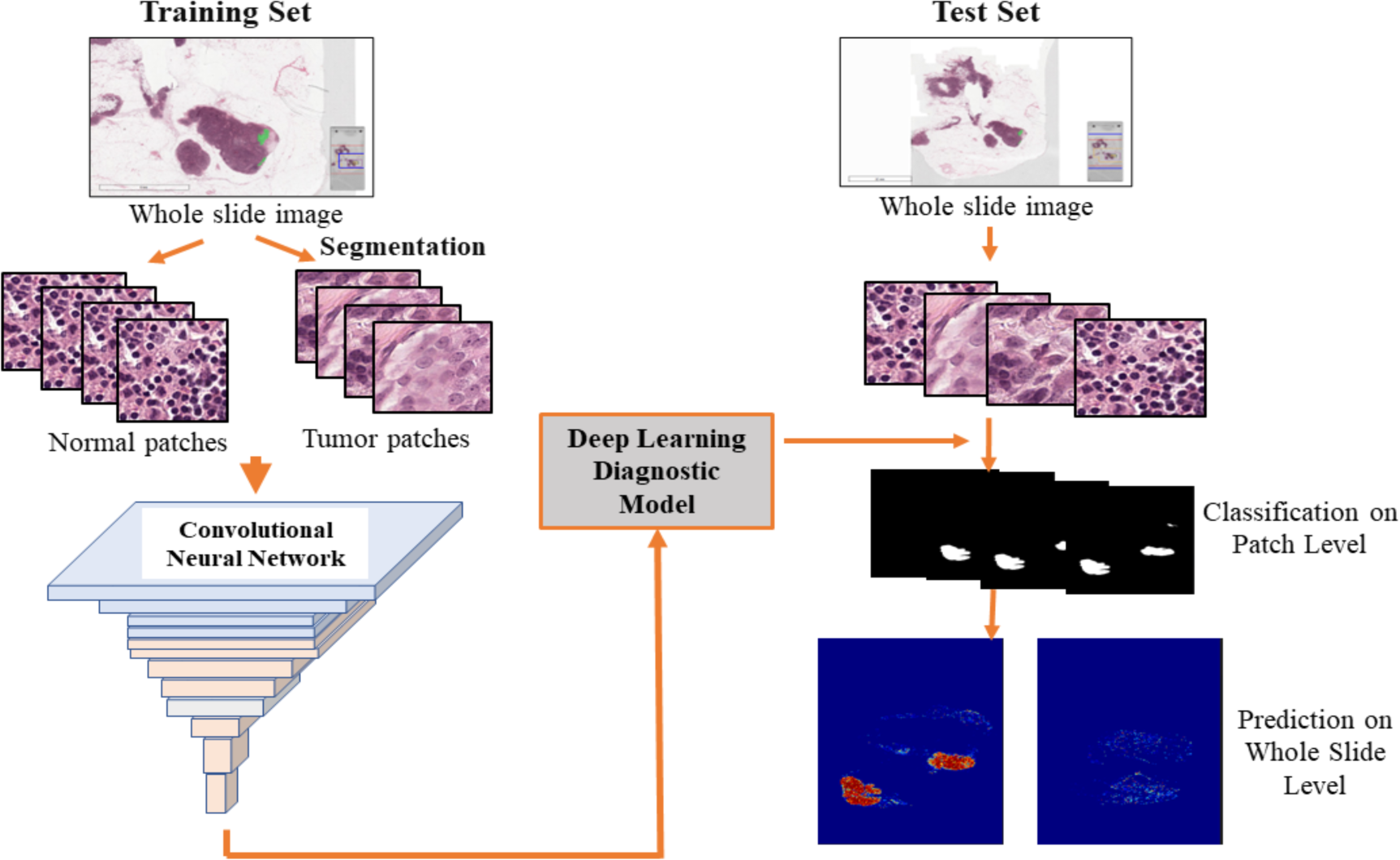
The Workflow of Deep Learning-Based Metastatic Breast Cancer Prediction

### 2.4. Federation of single-institution diagnostic systems

The federated learning-based system was implemented with the FederatedAveraging (FedAvg) algorithm ^[12]^ (Figure 3).

**Figure 3.**
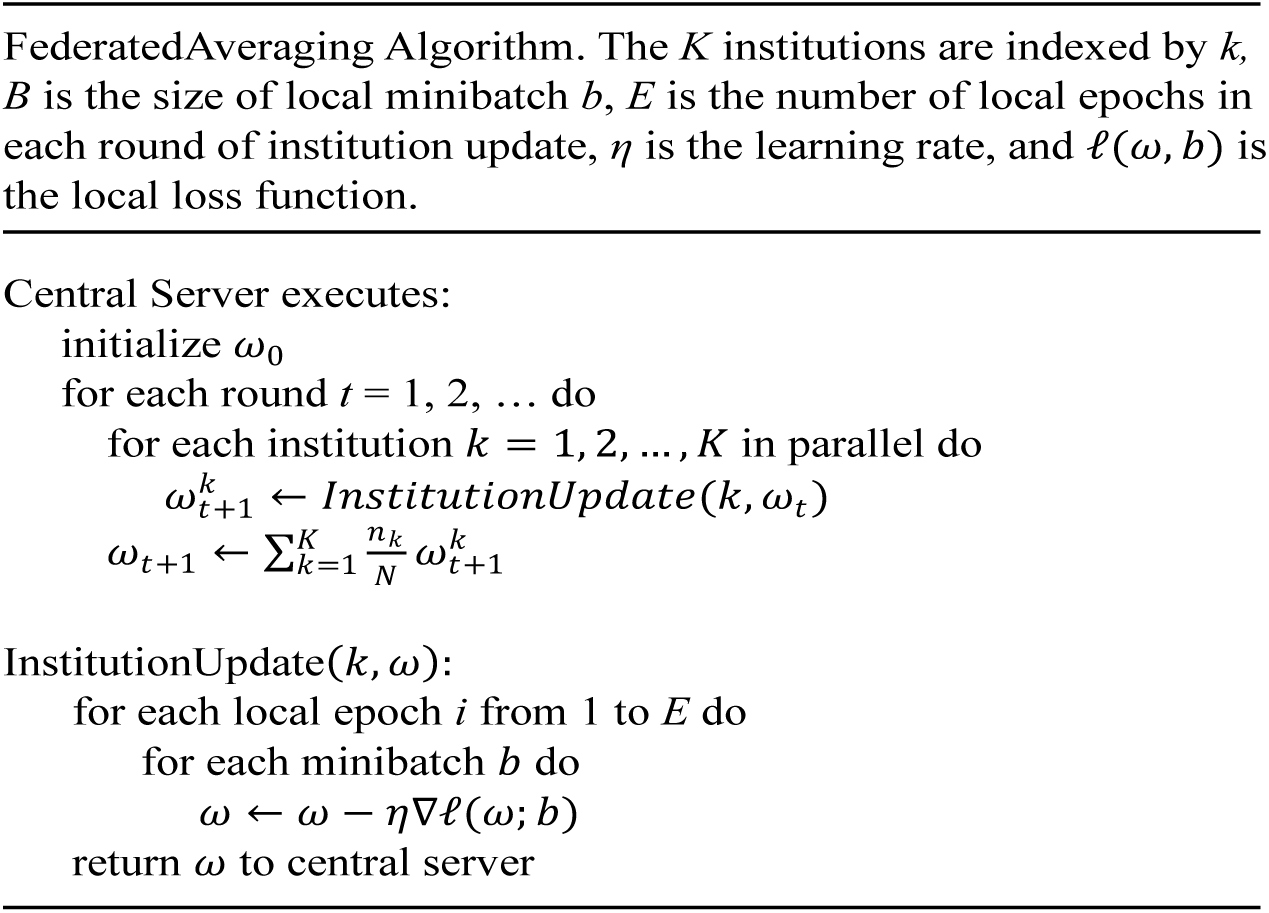
Pseudo-code of FederatedAveraging (FedAvg) algorithm ^[12]^

Where *η* is the learning rate, *n_k_* is the sample size of the training data set in k^th^ institute, and N is the total sample size combining all institutions. The federated averaging algorithm was integrated with the stochastic gradient descent (SGD) optimization at individual local institutions. In the federated collaborative diagnostic system, each collaborating institution *k* trained a diagnostic model of the same neural network architecture with learning rate *η* for one epoch and computed the average gradient ∇*f_k_*(*ω_t_*) on its local data at the current model *w_t_*. The central server aggregated the local gradients updated in this round of communication and applied the updates.

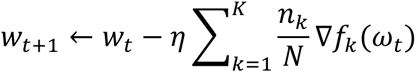

Computationally, this is equivalent to the central server taking a weighted average of the local models trained at all individual institutions. Each local model’s contribution was weighted based on its sample size. The FedAvg algorithm was implemented with the TensorFlow Federated API framework. Additionally, Mime Lite was explored to leverage a momentum parameter to decrease client drift and increase convergence rates. Hyperparameters were tuned for each model. The learning rate was adjusted to fine-tune convergence time and model performance. The batch size, number of clients, and client data volume were also adjusted for performance assessment. The number of training rounds (or epochs) was assessed in relation to convergence time.

### 2.5. Design of Simulation Experiments

We used simulation experiment to evaluate diagnostic performance of the federated collaborative diagnostic system for metastatic breast cancer.

We first assessed the ability of the federated learning-driven system in leveraging data from multiple institutions to improve diagnostic performance. This experiment simulated a scenario where the FL-based diagnostic system consisted of 4 simulated institutions (Figure 1B). For each training round, local models were trained at individual institutions, and the gradients from the local models were communicated to inform and update a federated model at a central server. The federated and local models were tested using an independent test set. Receiver operating characteristics (ROC) curve was used to evaluate diagnostic performance of the local models vs. the federated model.

We then explored whether the federated collaborative diagnostic system could improve accuracy of local diagnosis on lower-quality histopathological images. The lower image quality issue is a critical challenge for resource-constrained healthcare facilities in their effort of building high-performing models for diagnosis. We also used this experiment to further evaluate whether federated learning can improve model generalizability when data from multiple institutions are more heterogeneous in image quality. In this experiment, we simulated a scenario of 3 institutions, with institution C1 and C2 being healthcare facilities with adequate resources but institution C3 being an under-resourced facility. The experiment design is illustrated in Figure 4A. The image data at institutions C1 and C2 were of normal quality; the images at C3 were of lower quality (Figure 4B). The institution C3 images of lower quality were simulated by adjusting jpeg image quality to a random value between 6 and 9. A local model trained at a institution was tested on its own test dataset as well as the test sets at the other institutions. The federated model was also tested on the respective test set at each institution. The ROC curve was used for diagnostic performance assessment.

**Figure 4.**
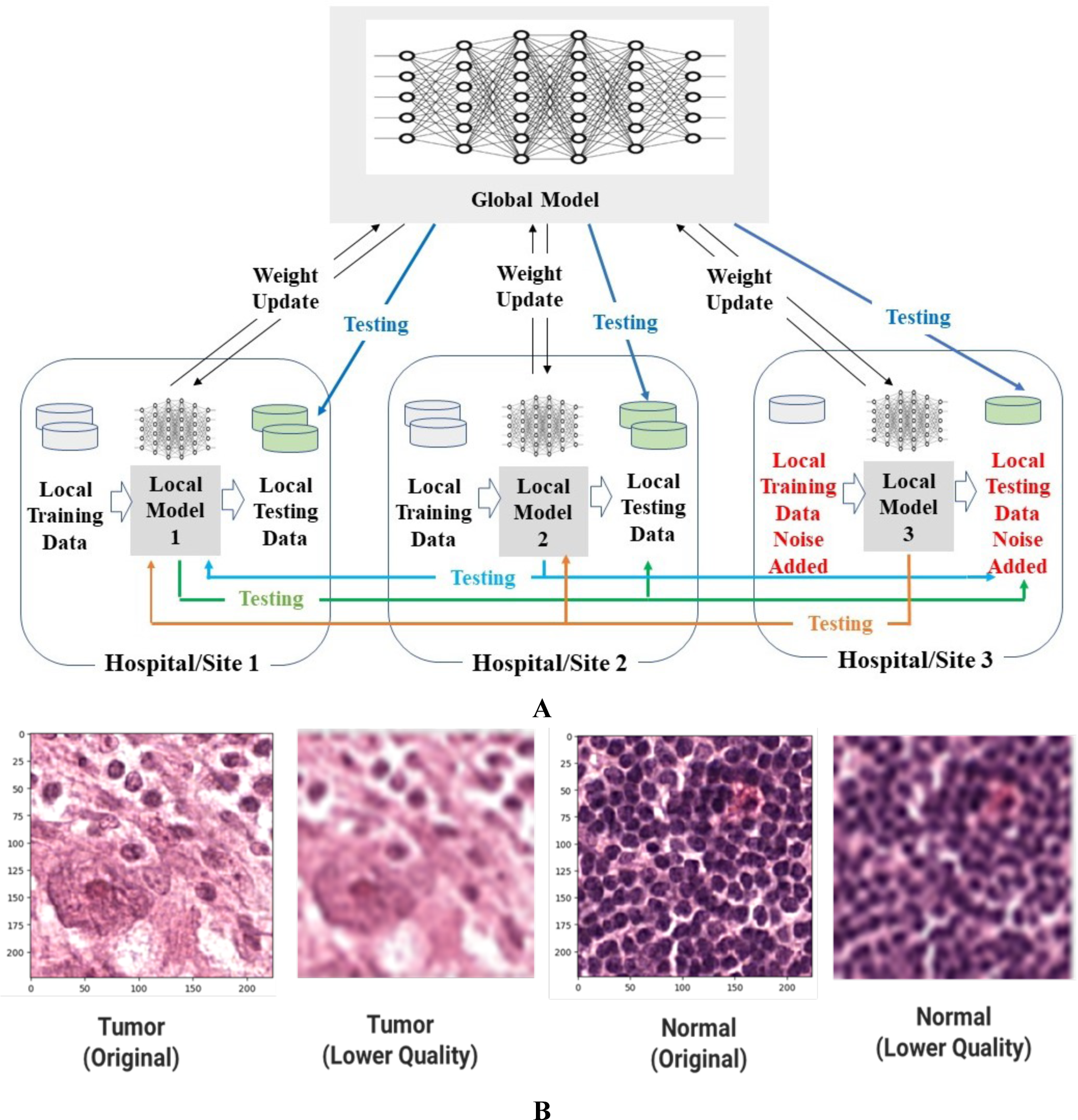
Experiment design to evaluate performance of federated model in improving diagnostic performance on lower quality images at a local site.

### 2.6. Datasets

The data consists of 222 whole slide images retrieved from the 2016 Camelyon ISBM challenge. The slides contain sentinel lymph node tissues extracted by the Radboud University Medical Center (Nijmegen, the Netherlands), as well as the University Medical Center Utrecht (Utrecht, the Netherlands).

## 3. Results

### 3.1. Federated Model Outperforms Local Models in Metastatic Breast Cancer Diagnosis

We evaluated diagnostic performance of federated model vs. local models for metastatic breast cancer diagnosis in a simulated scenario where a FL-based collaborative diagnostic system consisted of 4 local institutions (Figure 1B). The local models were trained, respectively, on the training dataset at each of the 4 institutions, and the gradients from the local models were communicated to inform and update a federated consensus model. Using an independent test set that was different than the training sets used at the 4 institutions, the federated model was evaluated against each of the four local models in diagnostic performance based on receiver operating characteristics (ROC) curve. As shown in Figure 5, the ROC AUC of the federated model (0.982) was higher than those of the local models (ROC AUC ranging from 0.831 to 0.940). The results suggest that the federated model can improve diagnostic performance for metastatic breast cancer by integrating local models from multiple institutions.

**Figure 5.**
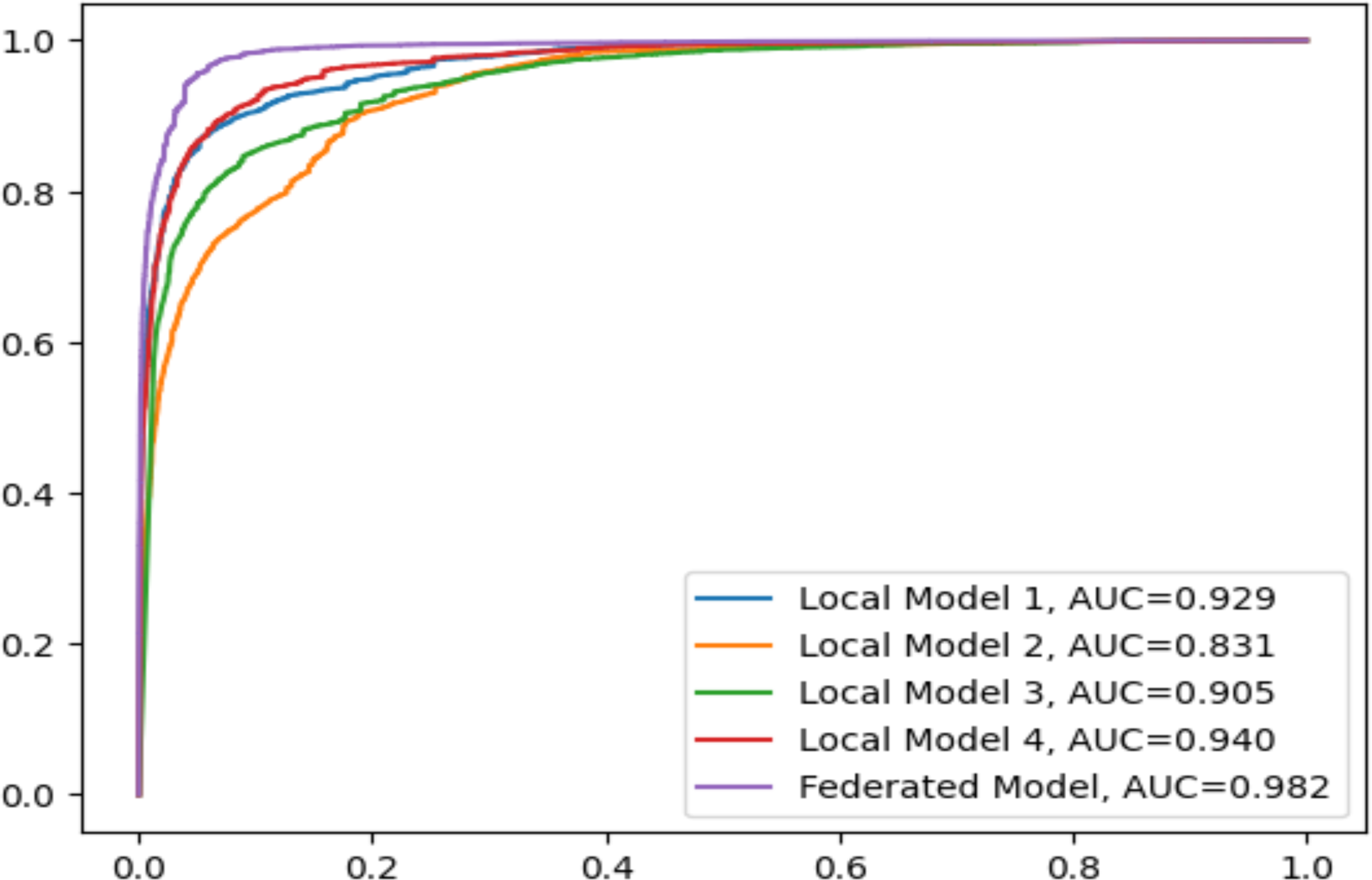
Receiver Operating Characteristics (ROC) curves of the federated model as compared with the local models from 4 collaborating sites.

### 3.2. Federated Learning Improves Diagnostic Model Generalizability

In the real-world scenarios, there exist variations in histopathological images among healthcare institutions due to differences in patient population, biopsy tissue processing, and histopathological slide preparation. Model generalizability is crucial for a diagnostic model to perform well for diverse patient populations and different tissue/image processing procedures. To evaluate generalizability of federated model, we conducted a simulation experiment on a federated collaborative diagnostic system that consisted of three institutions producing histopathological images of different quality. Among the three institutions in this experiment, the images generated at institution C3 were simulated to be of lower quality (Figure 4B). The dataset size of institution C3 was also reduced to reflect a shortage of image data at an under-resourced institution. The local models were trained and tested on the respective data at their own institutions. Further, the local models were tested on the test set at other institutions for generalizability assessment. Through the federated learning algorithm, the local models were aggregated to form a federated diagnostic model, which was then tested on the respective test set at each institution.

As shown in Figure 6, the federated model performed well for the test sets at all three institutions, with ROC AUC of 0.976, 0.980, and 0.985 for the test sets at institutions C1, C2, and C3, respectively. In comparison, all the local models showed a lower ROC AUC when applied to the test sets at other institutions. The result suggests that the federated model can improve model generalizability as compared with the local models.

**Figure 6.**
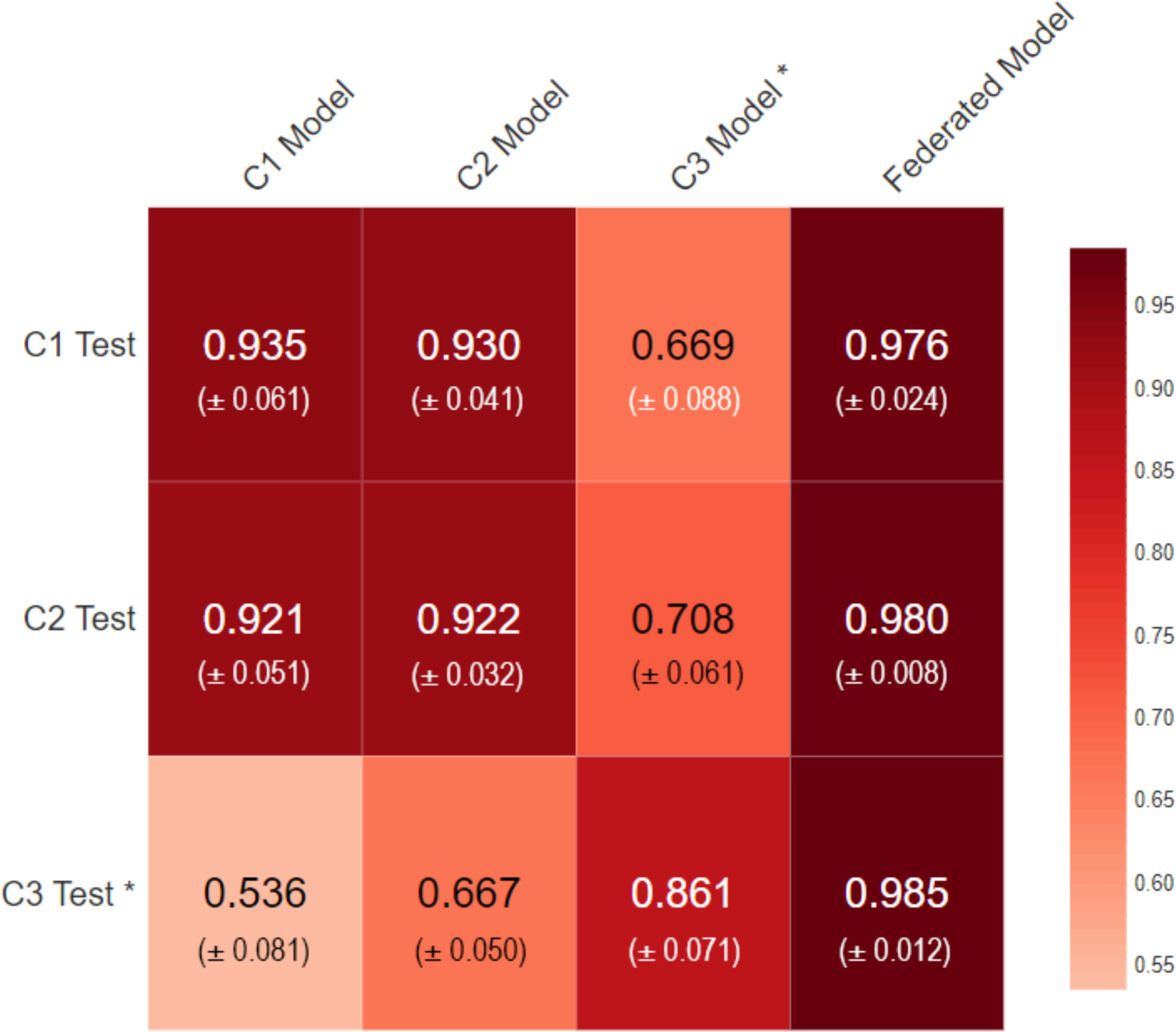
Diagnostic performance of the local models and federated model (average ROC AUC ± Standard Deviation): testing on respective test sets at individual institutions. * For institution 3, the histopathological images for training and testing were augmented to simulate images of lower quality.

### 3.3. Federated Model Improves Local Diagnostic Performance on Lower Quality Data

Lower-quality images hinder the effort in building high-performing diagnostic models in resource-constrained healthcare facilities. We used the experiment described above to evaluate whether a federated system can improve local diagnostic performance on lower-quality images. The local model trained at institution C3 had an ROC AUC of 0.861 on the test set at C3. Since both training and testing sets at C3 were of lower quality, the C3 model showed relatively lower diagnostic performance. Although the local C1 model had ROC AUC of 0.921 on the test set at C2 and the local C2 model had ROC AUC of 0.930 on the C1 test dataset, both local C1 and C2 models did not perform well on the C3 test set, with ROC AUC of 0.536 and 0.667 respectively. The results suggest that, while local models trained on normal quality data can perform well on test set with similar quality at a different institution, the local models might not be able to perform as well on the test set of lower quality because of limited model generalizability. However, the federated model achieved an ROC AUC of 0.985 on the test set at institution C3, which was a substantial increase of 12% as compared with the local model C3. The results demonstrate that the federated model can improve local diagnostic performance on lower quality images at an under-resourced institution.

### 3.4. Federated Model Improves Predictive Diagnosis on Unseen Whole Slide Images

To further evaluate federated model’s diagnostic performance on unseen whole slide images, visual comparison was conducted to examine diagnostic performance between the local models and the federated model on the unseen test set of WSIs, with the pathologist’s diagnosis as ground truth. As described in the previous section, the images at C3 were simulated to be of lower quality. Figure 7 shows the comparison for tumor case A. The boundaries of the cancerous regions predicted by the local model were not clearly defined. In comparison, the cancerous regions predicted by the federated model were better defined and more consistent with the pathologists’ diagnosis.

**Figure 7.**
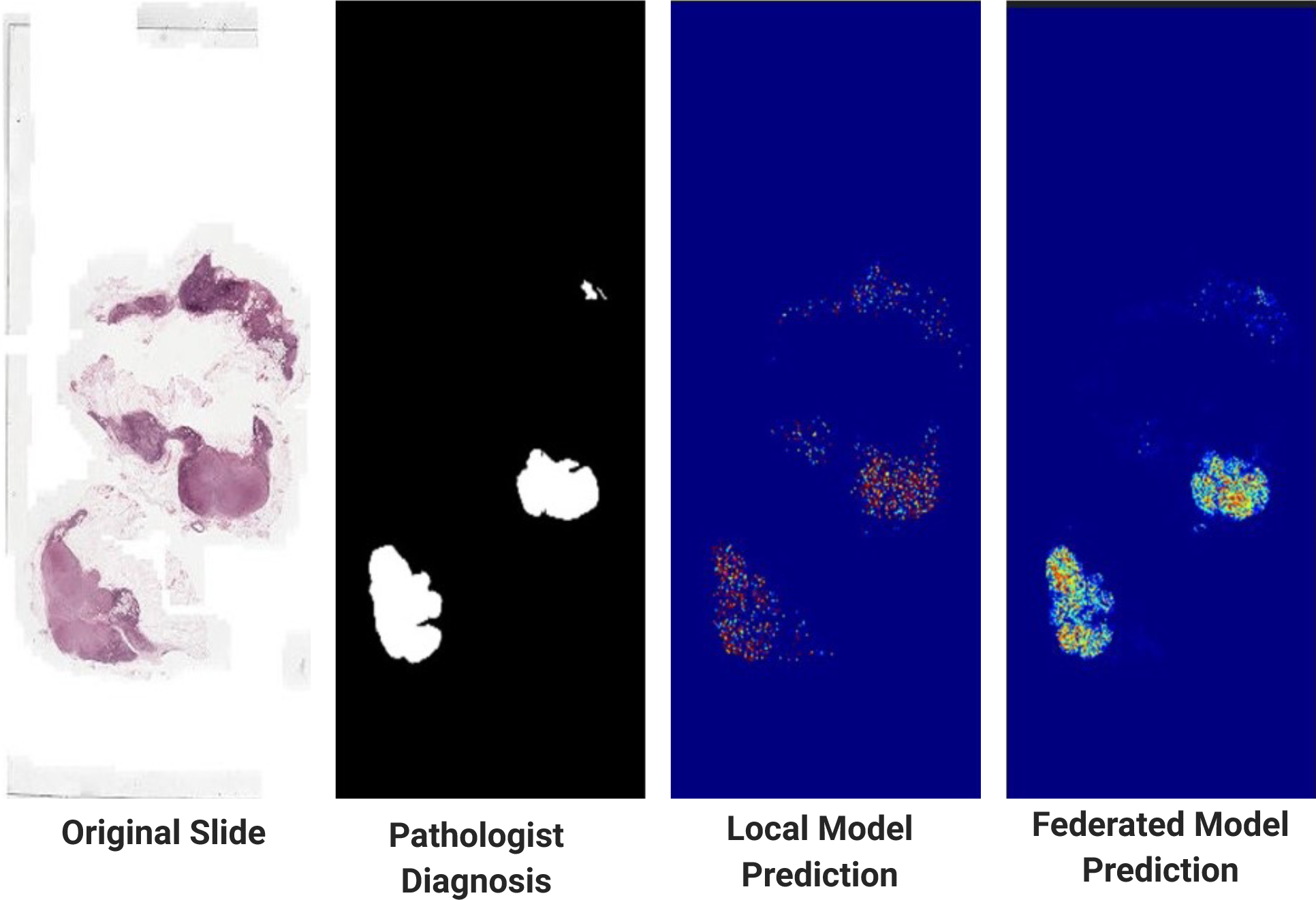
Tumor case A: comparison between the local model prediction and the federated model prediction on cancerous regions on WSI, with the pathologist’s diagnosis as ground truth

Figure 8 shows the comparison for a more challenging tumor case B. In this case, the cancerous regions were very small and embedded in a large area of normal cells. The local model cannot identify the small cancerous regions. In comparison, the federated model was able to identify the small cancerous regions. The boundaries were clearly defined and is consistent with the pathologists’ diagnosis. This comparison suggests that the federated model has a strong generalization and the ability to improve the identification of small cancerous regions on images of lower quality.

**Figure 8.**
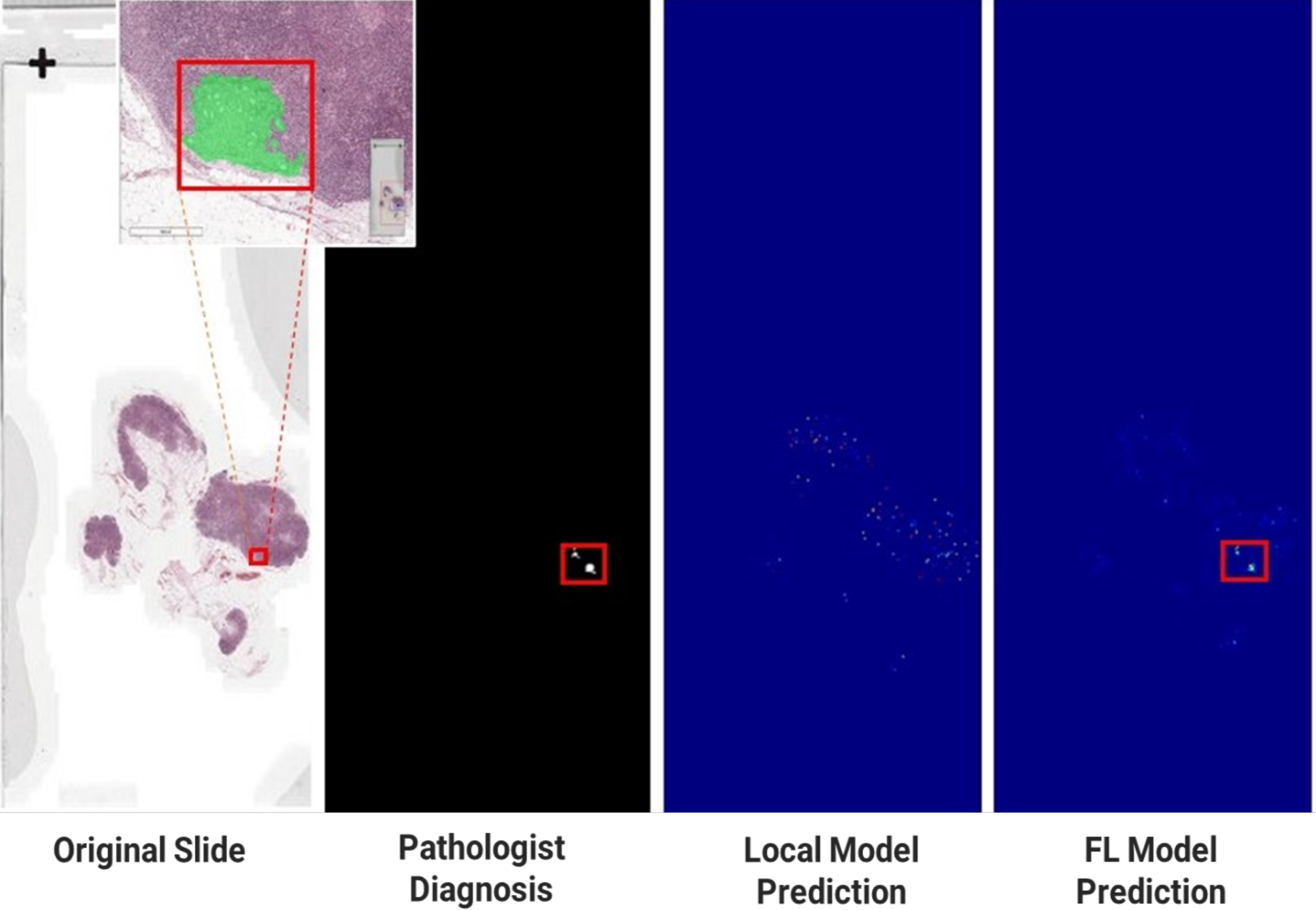
Tumor case B: comparison between the local model prediction and the federated model prediction on cancerous regions on WSI, with the pathologist’s diagnosis as ground truth

## 4. Discussion

In the development of deep learning methods for computer-aided image analysis for cancer diagnosis, it has been recognized that diagnostic model generalizability is largely dependent on size and diversity of training data. It was observed that diagnostical models trained from one healthcare institution may not perform equally well on the unseen data from different institutions ^[2]^. We also observed this issue in this study: the diagnostic models trained with normal data from institutions C1 and C2 did not perform equally well on the test set of lower quality images at institution C3 due to the local models’ limited generalizability. Therefore, building a generalizable diagnostic model requires access to rich and diverse sources of medical image data from multiple medical institutions. The current way for data aggregation is through direct transfer of medical data from multiple healthcare institutions, which is considered to be practically infeasible for real-world applications due to patient privacy, confidentiality, and legal obligations ^[10, 11]^. This challenge becomes more evident and severe for healthcare facilities in resource-constrained regions.

We propose a federated learning-driven collaborative diagnostic system for metastatic breast cancer in this study. Federated learning decouples model training from the need for direct medical data transfer, mitigating the potential risk for patient privacy and confidentiality. Further, this system can improve diagnostic accuracy and generalizability by leveraging knowledge derived from diverse data sources across multiple institutions. This study demonstrated that the federated diagnostic models performed consistently and substantially better than the local models. The improvement of model generalizability by federated learning may be a result of greater exposure of a federated model to data variations due to differences in biopsy sample acquisition, processing, equipment configuration, etc. among multiple institutions. The advantages of the federated learning-driven system can empower regional, national, and international healthcare institutions to collaborate on training and testing diagnostic models for metastatic breast cancer (Figure 9A). While the conceptual framework of such a collaborative diagnostic system is valid and technically feasible, more technical issues such as more robust privacy preserving solutions, client-server communication efficiency, etc. remain to be further explored.

**Figure 9.**
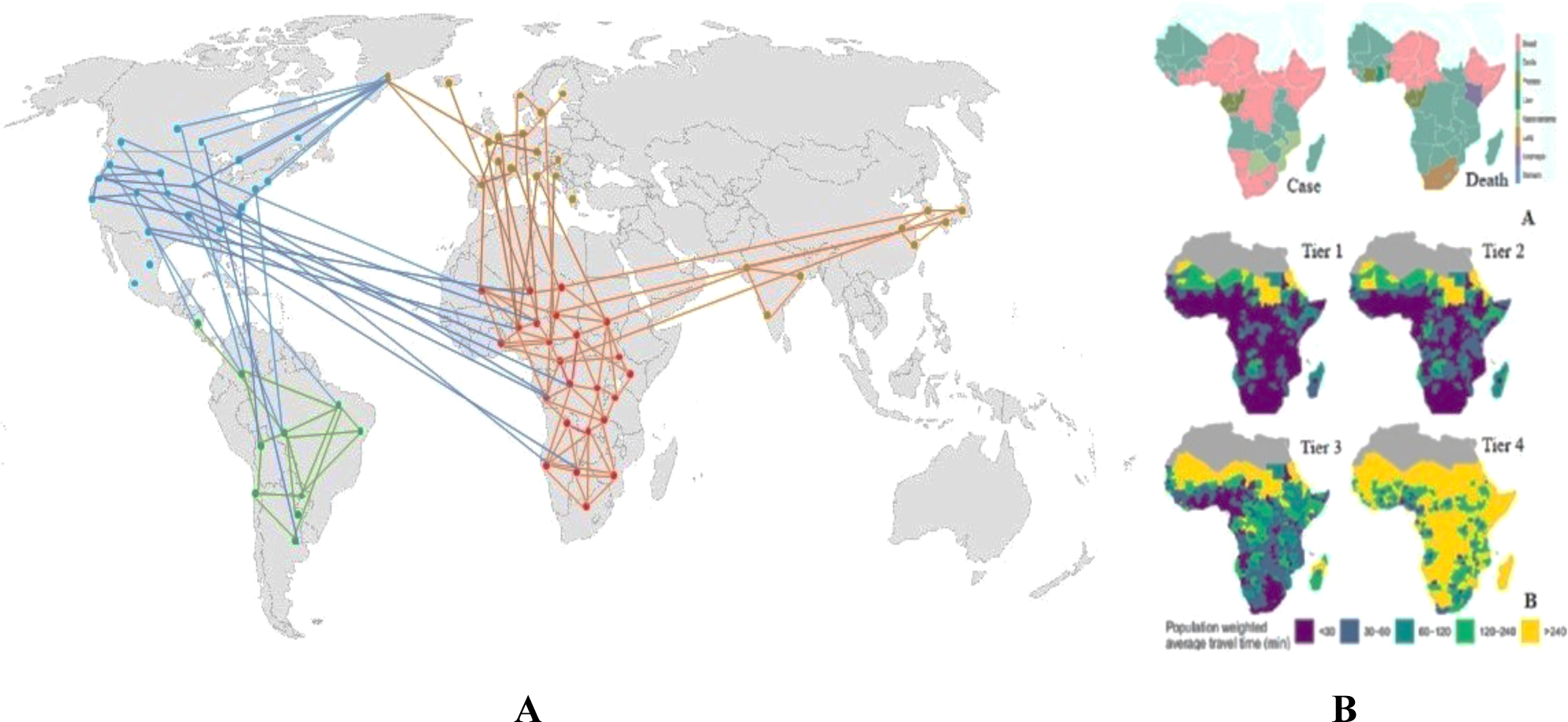
**A:** Federated learning-driven collaborative diagnostic system can be applied on a global scale to connect many healthcare institutions to improve diagnostic models while preserving patient privacy. **B:** Application of the system to connect all tiers (Tier 1 – 4) healthcare facilities (B) ^[21]^ to reduce the diagnosis delay in Sub-Saharan regions with high breast cancer cases and high mortality rates (A) ^[15]^.

The federated learning-driven collaborative diagnostic system provides an innovative approach to leveraging global data resources to address the public health challenges in metastatic breast cancer diagnosis faced by the developing countries. Breast cancer patients in sub-Saharan Africa, South Asia, and South America, suffer from the highest mortality rates in the world due to long diagnosis delays ^[8, 15, 22]^. In Northern sub-Saharan Africa, breast cancer has been the most common cause of cancer death, and healthcare resources are severely limited in this region ^[15, 21]^ (Figure 9B). A deep learning-based diagnostic system has been developed to address the disparity in breast cancer diagnosis, focusing on resource efficiency and mobile readiness ^[6]^.

However, its real-world application in the under-resourced regions still face a major challenge of severe shortages of high-quality medical image data needed for building high-performing diagnostic models. This study showed a dilemma in diagnostic model training under such a circumstance. At the simulated under-resourced institution C3, accuracy of the locally trained model was suboptimal because of the limited data volume and quality. On the other hand, the diagnostic model trained with a larger training set of normal quality images at other institutions (sites C1 and C2) underperformed on the unseen images at C3, suggesting limitations of deployment of an externally trained model for a local population. Federated learning provided an innovative solution to this problem. This study showed that the federated model achieved a ROC AUC of 0.985 on the test set at C3, an increase of 12% as compared with the local C3 model and an increase of about 30% as compared with the externally trained C1 and C2 models. The improvement of diagnostic performance by the federated model may be attributed to its ability in leveraging external information as well as adapting to local data.

Further, this study explored the potential of using federated learning to improve local diagnostic performance in an under-resourced healthcare institution through a collaborative diagnostic network. The under-resourced healthcare institutions are often located in remote, geographically isolated, and sparsely populated areas, e.g., northern sub-Saharan region (Figure 9B). Typically, these institutions face a severe shortage of trained pathologists. For instance, the average number of pathologists per head of population is 1 to 1,000,000 in sub-Saharan regions, compared with the ratio of 1 pathologist to 15,000–20,000 in the US and UK ^[7]^. This is further complicated by image quality issues due to lack of resources and training. In this study, the federated model substantially improved local diagnostic performance (measured by ROC AUC) by 12% on the lower quality images at C3. The visual comparisons have shown that the federated model can perform better than the local model on identifying cancerous regions, including the very small cancerous areas embedded in a large area of normal cells (Figure 7 and 8). This collaborative system presents a new opportunity to address the long delays in breast cancer diagnosis and consequent disparity in patient survival outcomes in developing countries.

## 5. Conclusion

In summary, this research has developed a federated learning-driven collaborative diagnostic system for metastatic breast cancer. This system preserves patient privacy by decoupling model training from the need for direct transfer of medical image data between healthcare institutions. Further, this system improves diagnostic model accuracy and generalizability by leveraging information derived from diverse data sources across multiple institutions. This study has demonstrated that this system has a great potential of improving local diagnostic performance on lower quality images at a resource-constrained institution. This research provides a new paradigm for metastatic breast cancer diagnosis – collaborative diagnostic model building and testing among multiple institutions around the world – to improve diagnostic accuracy and generalizability.

## Data Availability

All data produced are available online at https://camelyon16.grand-challenge.org/

## Authors’ contributions

W.G. conducted analysis and drafted the manuscript. D.W. and Y.H. provided advice on machine learning and statistical analysis. All authors reviewed and edited the manuscript.

## Availability of data and materials

The computational code used in this study will be made available upon request after the publication of this manuscript. Please contact W.G. for code.

## Declarations

### Ethics approval and consent to participate

Not Applicable

### Consent for publication

Not Applicable

